# Learning from pandemics: using extraordinary events can improve disease now-casting models

**DOI:** 10.1101/2021.01.18.21250056

**Authors:** Sara Mesquita, Cláudio Haupt Vieira, Lília Perfeito, Joana Gonçalves-Sá

**Affiliations:** Social Physics and Complexity Lab - SPAC, LIP, Avenida Prof. Gama Pinto, 1600-078 Lisboa, Portugal; Physics Department, Avenida Rovisco Pais, Instituto Superior Técnico, 1049-001, Lisboa, Portugal; Nova School of Business and Economics, Rua da Holanda, 2775-405 Carcavelos, Portugal; Instituto Gulbenkian de Ciência, Rua da Quinta Grande, 2780-156 Oeiras, Portugal

## Abstract

Online searches have been used to study different health-related behaviours, including monitoring disease outbreaks. An obvious caveat is that several reasons can motivate individuals to seek online information and models that are blind to people’s motivations are of limited use and can even mislead. This is particularly true during extraordinary public health crisis, such as the ongoing pandemic, when fear, curiosity and many other reasons can lead individuals to search for health-related information, masking the disease-driven searches. However, health crisis can also offer an opportunity to disentangle between different drivers and learn about human behavior. Here, we focus on the two pandemics of the 21st century (2009-H1N1 flu and Covid-19) and propose a methodology to discriminate between search patterns linked to general information seeking (media driven) and search patterns possibly more associated with actual infection (disease driven). We show that by learning from such pandemic periods, with high anxiety and media hype, it is possible to select online searches and improve model performance both in pandemic and seasonal settings. Moreover, and despite the common claim that more data is always better, our results indicate that lower volume of the right data can be better than including large volumes of apparently similar data, especially in the long run. Our work provides a general framework that can be applied beyond specific events and diseases, and argues that algorithms can be improved simply by using less (better) data. This has important consequences, for example, to solve the accuracy-explainability trade-off in machine-learning.

## Introduction

Infectious diseases pose great health risks to human populations worldwide. To mitigate these risks, public health institutions have set up surveillance systems that attempt to rapidly and accurately detect disease outbreaks. These systems typically include sentinel doctors and testing labs, and enable a timely response which can limit and even stop outbreaks. However, even when in place, detection and mitigation mechanisms can fail, leading to epidemics and, more rarely, pandemics, as we are currently experiencing. In fact, disease surveillance mechanisms that only rely on highly trained personnel, are typically expensive, limited, and slow. It has been extensively argued that these should be complemented with “Digital Era” tools, such as online information, mobility patterns, or digital contact-tracing^1–3^. Online behaviours, such as searches on Google, have proven to be very relevant tools, as health information seeking is a prevalent habit of online users^4^. This methodology has been applied to follow other epidemics, such as Dengue^5–7^, Avian Influenza^8^, and Zika surveillance^9^. In the case of Influenza, a very common infectious disease, the potential of online-based surveillance methods gained large support with the launch of Google Flu Trends (GFT), in 2008^10^. GFT attempted to predict the timing and magnitude of influenza activity by aggregating flu-related search trends and, contrary to traditional surveillance methods, provided reports in near real-time^11^, without the need for data on clinical visits and lab reports. More recently, many others have found strong evidence that the collective search activity of flu-infected individuals seeking health information online provides a representative signal of flu activity^12–16^. However, flu infection is not the sole (and perhaps not even the strongest) motivation for individuals to seek flu-related information online^17^. This is particularly true during extraordinary times, such as pandemics, when it is reasonable to expect individuals to have various degrees of interest, ranging from curiosity to fear, to actual disease^18^. In fact, the GFT model missed the first wave of the 2009 flu pandemic and overestimated the magnitude of the severe 2013 seasonal flu outbreak, in the USA^17,19^. This led many authors to suggest that high media activity can lead to abnormal Google search trends, possibly leading to estimation errors^17,20–24^. This “media effect” was also observed by others studying Zika^25–27^, and contributed to the disenchantment with the potential of such tools, particularly during such “extraordinary times”.

However, if we could decouple searches mostly driven by media, anxiety, or curiosity, from the ones related with actual disease, we could not only improve disease monitoring, we could also deepen our understanding of online human behavior. In the case of Google search trends, identifying what terms are more correlated with media exposure and reducing their influence in the model is crucial to correct past errors.

In this paper, we propose that the characteristics that make pandemics unique and hard to now-cast, such as media hype, can also be used as opportunities for two main reasons: 1) as pandemics tend to exacerbate behaviors, the noise (media) is of the same order of magnitude as the signal (cases), making it more visible, allowing us to discriminate between the two; and 2) because information seeking becomes less common as the pandemic progresses^18,28^ and these different dynamics can be used when selecting the search terms. In fact, instead of ignoring pandemic periods, studying what happens during the worst possible moment can help us understand which are the search-terms more associated with the disease and the ones that were prompted by media exposure. This solution might avoid over-fitting and enable the predictive model to be more robust over time, especially during seasonal events. Therefore, we focus on the only two XXI century WHO declared pandemics and aim at learning from pandemics to now-cast seasonal epidemics (or secondary waves of the same pandemic), and improving current models by incorporating insights from information-seeking behavior.

The first pandemic of the XXI century was caused by an Influenza A(H1N1)09pdm strain (pH1N1), which emerged in Mexico in February 2009^29^. By June 2009, pH1N1 had spread globally with around 30 000 confirmed cases in 74 countries. In most countries pH1N1 displayed a bi-phasic activity: a spring-summer wave and a fall-winter wave^30,31^. The fall-winter wave was overall more severe than the spring-summer wave as it coincided with the common flu season (in the Northern Hemisphere), that typically provides optimal conditions for flu transmission^32^. The pandemic was officially declared to be over in August 2010 and a total of 18 449 laboratory-confirmed pH1N1 attributable deaths were counted (WHO, 2009). This number was later revised and pH1N1 associated mortality is now believed to have been 15 times higher than the original official number^33^. The second pandemic of this century, was caused by the SARS-CoV-2 virus, first identified in the last day of 2019 in the Chinese province of Wuhan. To date, Covid-19 has infected more than 78 million people and killed more than 1,7 million people worldwide.

Both Covid-19 and influenza viruses cause respiratory diseases with manifestations ranging from asymptomatic or mild to severe disease and death. They share a range of symptoms and trigger similar public health measures due to common transmission mechanisms. Both pandemics have a led to a great surge in media reports and public attention across many platforms, from traditional to online social media. However, there are several differences between the two pandemics: there is still a lot of uncertainty and lack of knowledge surrounding the SARS-CoV-2 virus, including its lethality (although it is certain to be higher than the flu for older age-groups), whether it displays seasonal behaviour, its transmission frequency and patterns, whether infection confers lasting immunity, or what are its long-term health effects, respiratory or not^34–38^. Moreover, the Covid-19 pandemic led to unique public health measures and what might be considered the largest lockdown in history, with authorities implementing several preventive measures from social distancing to isolating entire countries. These restrictions have been instrumental in reducing the impact of the pandemic, but most decision-makers acknowledged the need to loosen the confinement measures. In the interest of economic and social needs, several countries re-opened schools and businesses, and many experienced surges in cases and deaths^39^, often referred to as second and even third waves. At this point, and as vaccines start to be distributed mostly in developed countries, all tools that can help us in identifying outbreaks are of utmost importance and different countries are deploying different measures such as conditional movement and contact tracing apps.

For all these reasons, improving fast, online surveillance is even more crucial now than it was in 2009, and there are already several studies on using online data to explain and forecast Covid-19 dynamics^40–45^. However, and despite its potential, separating what is media hype from reporting of actual disease cases (be it on Google, Facebook, or any other platform), and understanding their impact on collective attention, has been considered a huge challenge. One of the main reasons is that the patterns are intertwined with the actual spread of a disease within a population. Therefore, we learn from the 2009 flu pandemic and propose a system to reduce the signal to noise ratio on online-searches and now-cast the current Covid-19 pandemic. The 2009 influenza offers a great case study as it was extensively researched: precise signals of pandemic flu infections were obtained through large-scale laboratory confirmations^46^, several studies analyzed the media’s behaviour during the pandemic^47–49^, including the collection of news pieces and news counts, and as the pandemic emerged at a period of widespread Internet usage^50^, several online datasets are available (including the collective behaviour of millions of users through their search trends on Google). Building on these datasets and by adding insights from human behaviour, we apply our framework to the current Covid-19 pandemic and provide a robust and possibly generalizable system.

## Results

### Dynamics of media reports and online searches do not match disease cases

Improving signal (disease) to noise ratio is fundamental in disease surveillance. As extraordinary events, such as pandemics, tend to become the dominant story nearly everywhere, fear and curiosity can increase and so do searches for information.

First we asked whether there is a correspondence between the number of cases (for both the 2009 flu and Covid-19), media reports, and searches on Google for disease-related terms (flu and Covid-19, respectively). We focused on the US in the case of the 2009 flu and Spain during the Covid-19 pandemic. These are countries that had a large number of cases, good data availability and, in the case of Spain, already a strong second Covid-19 wave, as detailed in the methods. Figure 1 shows the number of confirmed infections, news mentions and GT searches in the United States for the 2009 pandemic (a) and in Spain for the current one (b). Since news now travel faster than pathogenic agents, the news peak for the 09 flu pandemic (figure 1a) had its peak on the last week of April, while the first peak in cases happened later, at the end of June. More relevant is that by the time H1N1 infections had its highest peak in the US (in October/November, during regular flu season), the frequency of online searches for “flu” and news mentions had significantly reduced. In the case of the Covid-19 pandemic (figure 1b), the early news mentions began in late 2019 when the disease was identified in China, but the first cases in Spain were only identified in February 2020 (for a similar analysis on the US case see the supplementary materials). As observed before, there was a disconnect between the intensity of the disease and both its visibility in media and the volume of Google searches^17,19^, raising the important question of whether we can discriminate between different drivers of online searches.

**Figure 1.**
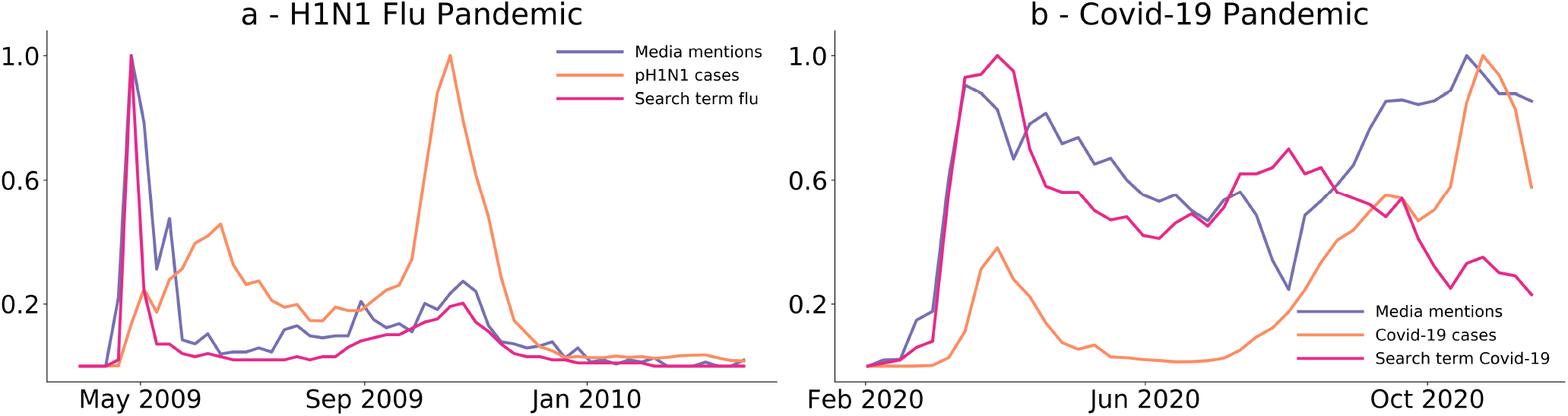
Flu and Covid-19 cases during the 2009 and 2020 pandemics in US (a) and Spain (b), respectively. **a -** Normalized weekly cases of flu (orange), media mentions (purple), and Google-trends searches for the term “flu” (pink) in the United States of America from March 2009 to March 2010. **b -** Normalized weekly cases of Covid-19 (orange), media mentions (purple), and Google-trends searches for the term “Covid-19” (pink) between February and November 2020. All datasets are normalized to their highest value in the period. We can see a quick increase in media activity in both situations that precedes the number of cases of infection. In both panels, searches for the terms ‘flu’ or ‘Covid-19’, display a pattern more similar to the media activity trend (Pearson correlation between the search term and media of 0.85 for the flu pandemic and 0.44 for Covid-19 pandemic, compared to 0.27 and −0.03 between the search term and cases of infection, respectively).

### Online searches have different patterns

Given that the searches for “flu” and “Covid-19” do not closely follow the variation in the number of confirmed cases, we asked if we could identify particular search terms, with higher correlation with the disease progression. We started by selecting a large number of search terms, related to each disease (see supplementary materials for the full list), all of which could be *a priori* considered useful for now-casting. Using hierarchical clustering, we identified three distinct clusters in both the 2009 flu and COVID-19 (2a and 2d). Figures 2b and 2e show the centroids of each cluster, revealing the existence of different dynamics. In the case of the flu in the US, one cluster has a strong peak in the second half of 2009, another has the strongest (almost unique) peak in the first half, and a third cluster has much less clearly defined peaks (figure 2b). The first cluster (orange) shows a strong correlation with the number of pH1N1 confirmed cases (*r* = 0.78, *p* = 4*×* 10^*−*16^) and a lower correlation with media (*r* = 0.60, *p* = 2*×* 10^*−*8^), while the second cluster (purple) has the opposite trend (figure 2c, *r* = 0.16, *p* = 0.2 with pH1N1 cases and *r* = 0.83, *p* = 3 *×*10^*−*20^ with media). The third has an intermediate correlation with the flu cases and poor with the media reports. As an additional test, we asked whether there was evidence that cases or media preceded any of the clusters. We performed a Granger causality test and show that that media precedes cluster 2 but not cluster 1 (supplementary materials). Neither cases nor media showed significant results for clusters 1 or 3. The grouping of the search terms is not intuitive from their meaning. Interestingly, there is no clear pattern on the search-terms that could have indicated that some would be more correlated with cases or media attention. For example, symptoms such as ‘fever’ or ‘cough’, appear on cluster 3, together with ’Guillaume-Barré syndrome’ and disinfectant’, while cluster 1 contains ’vaccine’ and ’treatment’ along with the strain of the virus and ’hand sanitizer’.

**Figure 2.**
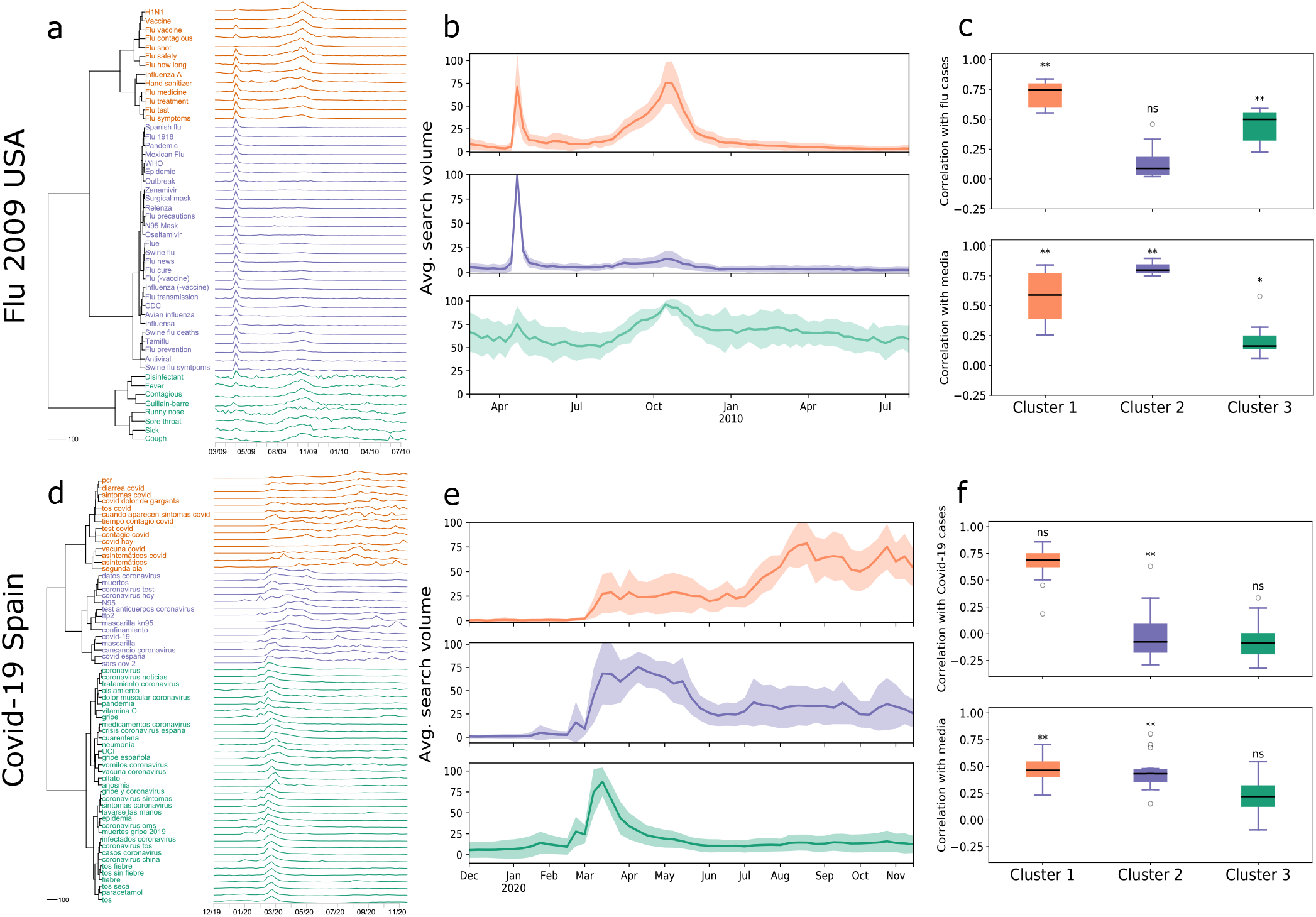
Different patterns of searches during pandemics. Top panels refer to the 2009 flu pandemic in the USA, bottom panels refer to the COVID-19 pandemic in Spain. **a -** Dendrogram summarizing the hierarchical clustering of Google Trends search terms for the flu pandemic in US. Three clusters are very salient. **b -** Centroid and standard deviation over time for each cluster. The cluster colors correspond to the clusters in a. **c -** Pearson correlation between the cluster centroid and either the flu cases (top) or the media mentions (bottom).* denotes 0.01 < *p-value* < 0.05, ** denotes *p-value* < 0.001, and *ns* a non significant *p-value*. **d -** Dendrogram summarizing the hierarchical clustering of Google Trends search terms for Covid-19 in Spain. **e -** Shows the centroid and standard deviation over time for each cluster. The cluster colors correspond to the clusters in d. **f -** Pearson correlation between the cluster centroid and either the Covid-19 cases (top) or the media mentions (bottom).

In the case of Covid-19, the clusters are not so well defined, as shown by the smaller relative length of the internal branches of the clustering dendrogram (figure 2d). This is likely due to a) the smaller time-frame considered (roughly half of that of H1N1 - figure 1) b), the lower search volume, explained by the much smaller population of Spain when compared to the US, and c) the real-time nature of the analysis. Still, we could identify three clear clusters and a very similar pattern (figure 2e): the first cluster (again orange) shows two broad peaks, the second larger than the first. The second cluster (purple) shows a clear first plateau, between March and May 2020, and the third cluster (green) a much sharper peak, encompassing little over one month. When we repeated the correlation analysis, we again identified a cluster (C1, orange) that strongly correlates with the number of cases (*r* = 0.71, *p* = 8 *×* 10^*−*6^) but less with the media (*r* = 0.52, *p* = 0.003), and a cluster (C2, purple) with the opposite pattern (a correlation with cases of *r* = 0.13, *p* = 0.45 and with media of *r* = 0.71, *p* = 2 *×* 10^*−*6^) (figure 2f). Cluster 3 (green) correlates poorly with both the number of confirmed cases and media attention. Thus, and despite the strong entanglement and time-coincidence between the cases and the media, particularly in the case of the current pandemic, these results show that 1) not all pandemic-related search trends show the same patterns, and 2) some of the patterns may be driven by media attention whereas others by the number of cases.

### Pandemic search-terms can be used to improve seasonal forecasting

That very similar search-terms display such different time patterns is interesting in itself but only useful if they have predictive power. Therefore, we asked whether the search terms identified as correlating with the number of confirmed cases (during a pandemic) could be used to forecast seasonal epidemics. The rationale is that if we can reduce the noise caused by the media coverage and identify the terms that are more resilient to outside factors, we can make seasonal forecasting more robust. Therefore, our goal was not to devise the best possible model, but rather to test whether particular search terms perform better than others. To do this, we took advantage of extensively available seasonal flu data and chose two simple models: a linear regression and the non-linear random forest (details in the Methods). We then tested the predictive power of the models when we used all search terms from figure 2A (that we call “All data”) or just the terms from the identified clusters in figure 2b. For both models and all dataset variations, we used three years of data to predict the fourth and assessed the performance of the model only on the prediction season (see Methods for details). Figure 3 and table 1 show the performance of the two models (Figure 3a and 3b) measured by the root-mean-square error (RME) and the coefficient of determination, R^2^. In general, both models perform similarly, with a mean R^2^ above 0.7. In both cases, using all data (pink line) is not better than just using the terms more correlated with the number of cases during the pandemic (cluster 1, orange line), and on average cluster 1 performs better than all terms in both the linear regression (R^2^ = 0.81 for cluster 1 *vs* R^2^ = 0.71 for all data) and random forest(R^2^ = 0.86 for cluster 1 *vs* R^2^ = 0.81 for all data). It can also be observed that cluster 1 terms (orange) tend to have a more consistent performance (shown by the smaller standard deviation: 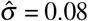 for cluster 1 in the case of linear regression and 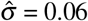 for random forest *vs* 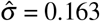 in the case of linear regression and 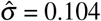 for random forest when considering all data).

**Table 1.**
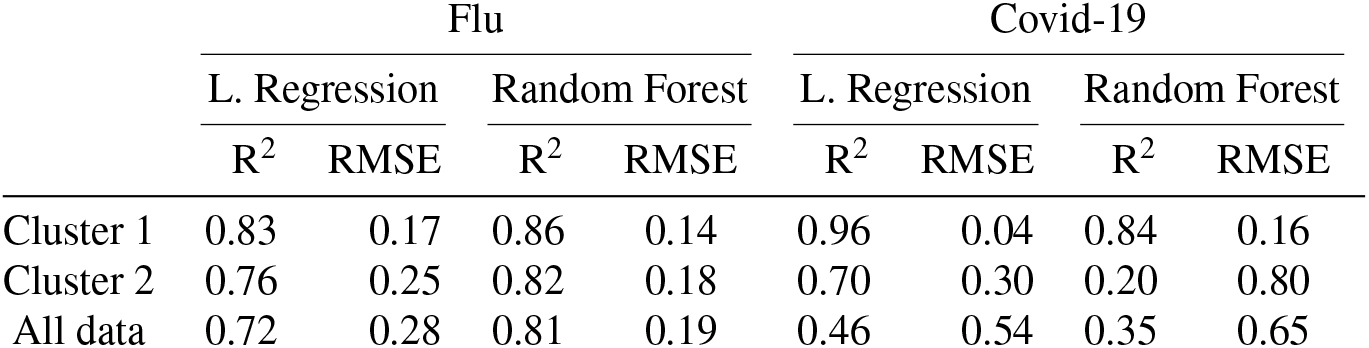
Model results for both pandemics.

**Figure 3.**
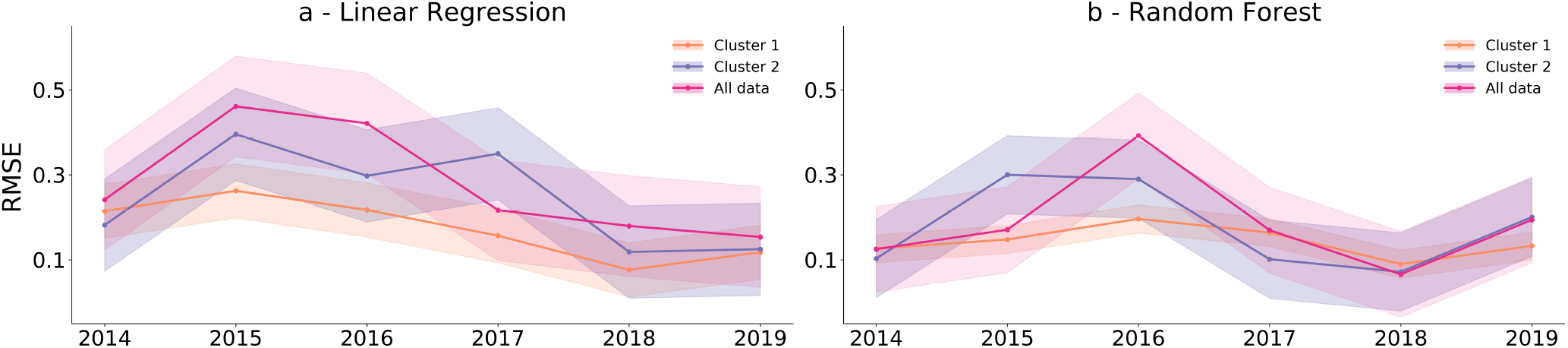
Performance comparison of model predictions for the flu pandemic. **a** shows the mean squared error for the linear regression and **b** for the Random Forest model. Both use Google search terms from figure 2a as independent variables to predict the seasonal flu cases between 2014 and 2019. Each dot represents the squared difference between the prediction and the empirical data, averaged over one season. Cluster 1 (orange) shows better results in almost all seasons and has a smaller standard deviation (shaded area) when compared to cluster 2 (purple) or all data (pink). In both cases, three years were used as training and the models were tested on the following year, in a sliding window process.

It is important to note that some of the features from clusters 2 and 3 might be better local predictors, and that can explain the performance of the models when using all search terms, but overall, using only the pre-identified terms of cluster 1 is better.

This indicates that 1) insights from pandemics can be used in seasonal forecasting models, and 2) refining the search-term selection, by selecting the ones less sensitive to media hype, might reduce over-fitting and improve model robustness.

### Improving a model for Covid-19

We then asked whether these results could be used in the current pandemic. This is a more challenging setting for several reasons: first, the data is arriving in close to real-time and with varying quality (the number of tests, the criteria for testing, and the reporting formats have been changing with time, even for the same country); second, there is no indication that Covid-19 might become a seasonal disease and the periodicity of new outbreaks, if any, remains unknown; third, reporting is now happening in many different online platforms, at an even faster pace than in 2009, and more importantly fourth, we do not have a large number of past seasons to train our models on. Still, we employed a similar approach to test whether the rationale of the flu pandemic could be applied to Covid-19. The US pandemic situation has been particular, with different states having widely different infection rates and risk levels^51^. Also, at the time of this study, there were no states with clear strong second waves or evidence of seasonality. Therefore, we focused on Spain, one of the first countries to have a clear and strong second wave and trained the models on the first (February-June) wave to try to now-cast the second (June-November) wave. Still, data for the US can be found in the supplementary materials with results very consistent with what we observed in the case of Spain. Figure 4 shows that, again, using only the features from cluster 1 (orange) offers a much better prediction than using the search-terms from clusters 2 (purple) or 3 (supplementary materials), despite the fact that cluster 1 has a much smaller number of terms.

**Figure 4.**
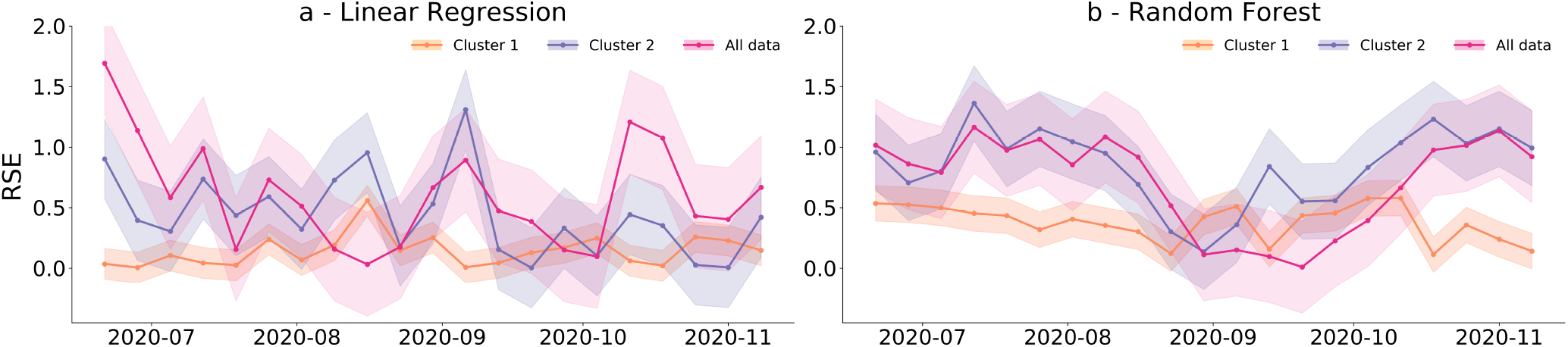
Performance comparison of model predictions for Covid-19. **a** shows the mean squared error for the linear regression and **b** for the random forest model. Both use Google search terms from figure 2d as independent variables to predict the second wave (June to November) of Covid-19 in Spain. Each dot shows the squared difference between the prediction and the empirical data in each week. Cluster 1 (orange) presents better results in almost all seasons and has a smaller standard deviation (shaded area) when compared to cluster 2 (purple) or all data (pink). In both cases, the first wave was used to train the model.

The result is particularly striking in the case of the random forest (figure 4b, compare pink and orange). These results further support the idea that by selecting online data, using a semi-manual approach, it is possible to improve disease now-casting.

## Discussion

In the past, the inclusion of online data in surveillance systems has both improved the disease prediction ability over traditional syndromic surveillance systems, while also showing some very obvious caveats. Online-data based surveillance systems have many limitations and challenges, including noisy data, demographic bias, privacy issues, and, often, very limited prediction power. Previous approaches have assumed that if a search-term is a good predictor of cases in one year, it will be a good predictor in the following years^11,52^, when in fact, search terms may be associated with both cases and media hype in a particular year, but soon loose association with one or the other (especially when media interest fades). Moreover, and taking into consideration that these approaches often use a single explanatory-variable, meaning the model ignores the variability in individual search query tendencies over time, it can happen that terms highly correlated with disease cases in a certain moment can be highly correlated with media reports as well, but over time some might lose their association with one or another. However, and despite the described limitations, there are several successful examples of using online behaviour as a proxy for “real-world” behaviour in disease settings and it is increasingly clear that such data can offer insights not limited to disease forecasting^16,53–56^.

Pandemics have been particularly ignored in digital now-casting because they represent (hopefully) rare events when people’s behaviour changes, making forecasting even more challenging. A large part of these behavioural changes is driven by the excess media attention: people become curious and possibly afraid, and start looking for more (different?) information. This is in contrast with seasonal outbreaks where there isn’t so much relative media attention, there is more common knowledge, and people’s online searches might be primarily driven by actual disease. In general, the notions that online search-data is too noisy and that the models used have limited prediction power have led people to try to increase the type and quantity of data, or to build more complex models. However, we argue that this tension, between using the large potential of online data and the so-called “data hubris”, can be balanced in the opposite direction, by including behavioural knowledge and human curation, to reduce the amount of data required, while keeping the models simple and explainable.

In this study, we applied this approach to two pandemics and showed that, contrary to general arguments of “more data trumps smarter algorithms”^57^ we can use such extraordinary events to improve seasonal forecasting, and argue that lowering the volume of data can reduce over-fitting while maintaining the quality of the predictions. This was done by actively discriminating between search queries that are very sensitive to media from queries possibly more driven by symptoms. Our approach combines elements of human curation and blind statistical inference. On the one hand our initial term list is based on knowledge of the disease. On the other, the clustering algorithm is blind to the actual meaning of the terms. This leads to unusual term-pairings such as the fact that “oseltamivir” (cluster 2), a drug used to treat flu is separate from “flu treatment” (cluster 1). We can explain this separation by considering that the media is more likely to mention the name of the drug, but that sick people might not remember it. However *a priori* we might not think this distinction was important. Finally, the choice of the best cluster is again based on human curation by looking at the correlations with media and cases, which we postulate are the main drivers behind search queries. In many general now-casting problems, a similar semi-automated approach is probably more fruitful than a fully automated, data-hungry methodology. This approach can also be particularly useful in countries where data is sparse or suffers from significant bias or delays. Even within Europe, data collection and reporting have been inconsistent, limiting global epidemiological analysis^57^. Methods as the one we describe here cannot replace the need for strong, centralized, data collection systems (through the European, American or other CDCs) but might help to fill existing gaps, while surveillance networks are built or reinforced.

In addition to improving now-casting models, finding different search patterns in Google Trends can offer insights into the behaviours of internet users. Specifically, by clustering search trends on a topic we can ask whether there are different motivations behind them. If there are hypotheses about what those motivations are, they can also be tested by correlating with centroids as we do here. For example, the search terms from the media-related clusters (clusters 2) could be further analyzed to discriminate which terms are more often found in newspapers versus television, offering insight into the preferred news media. This methodology opens new doorways into connecting online and offline behaviour.

Overall, we add to the ongoing work on using digital tools and online data to improve disease monitoring and propose a new tool to now-cast infectious diseases, combining statistical tools and human curation, that can prove useful in the monitoring of the current and future pandemics and epidemics.

## Methods

### Data and Sources

#### Selected countries and time period

Data for the 2009 pandemic was collected for the USA, from March 2009 to August 2019, as it offered reliable data on a large number of people. This was not possible for Covid-19 as this pandemic is reaching different states at different times and second or third waves are mostly caused by surges in new states than as a nation-wide, simultaneous epidemic. Still, supplemental text shows that three clusters are observed, one more correlated with cases than the rest. Data for the Covid-19 pandemic was collected for Spain, from January 2^nd^ to November 15^th^ 2020, as it was the country with highest number of reliable second-wave cases, offering at least one training and one testing period.

#### Google search trends

Data from Google search trends (GT)^58^ was extracted from the United States and Spain both for flu and Covid-19 pandemics, through the GT API. It provides a normalized number of queries for a given keyword, time and country^11,59^. Search terms were selected to cover various aspects of pandemic and seasonal flu, and Covid-19, such as symptoms, antivirals, personal care, institutions and pandemic circumstantial terms. This was done with the help of “related queries” option that Google Trends provides, returning what people also search for when they search for a specific term. Terms that contained many “zeros” interspersed with high values were indicative of low search volume and were removed. In the end we had 49 flu-related weekly search trends in the United States and 63 Covid-related terms in Spain. Time periods were December 2019 to September 2020 in the case of Spain and September 2009 to September 2019, in the case of the USA, to cover pre-pandemic, pandemic and post-pandemic periods. In the case of the US flu pandemic, search-terms were extracted for each season separately, with a season being defined as going from September 1^st^ to October 1^st^ the following year. GT time series were extracted in September 2020 in the case of Spain, and July 2020 in the case of the US. Data was binned in a weekly resolution, to match that of reported cases and remove daily variation. Both word lists are reported in the supplemental text.

#### News media

The pandemic flu, United States media dataset contains the weekly count of both TV news broadcast and print media, that mentioned “flu” or “influenza”. It includes NBC, CBS, CNN, FOX and MSNBC networks, obtained from the Vanderbilt Television News Archive^60^, and The New York Times, from the NYT API (https://developer.nytimes.com/). The Covid-19 media dataset, for both the USA and Spain was obtained through Media Cloud^61^, an online open-source platform containing extensive global news corpus starting in 2011. The query “Covid-19 OR Coronavirus” was used to track media coverage of the pandemic over time. It aggregated articles that had 1 keyword, the other or both. For the case of the US, we searched the collection “United States -National” (#34412234) and “United States - State & Local” (#38379429), which includes 271 national and 10,457 local media sources, respectively. For Spain we used collection “Spain - National” (#34412356) which includes 469 media sources, and Spain - State & Local(#38002034), including 390 media sources.

#### Infectious Disease Data

Data of confirmed infections from both pH1N1 and SARS-CoV-2 are publicly available. For US pH1N1 cases were extracted from the CDC’s National Respiratory and Enteric Virus Surveillance System^62^. In the case of Covid-19 in the US, data from national and state-level cases were extracted ECDC’s Our World in Data^63^ and from New York Times^64^, respectively, in August 2020. In the case of Covid-19 in Spain, data was obtained from the WHO^39^.

### Analysis

#### Hierarchical clustering

Google search terms were independently extracted from Google Trends^65^. While all search queries include a 100, not all include a zero (if there were no weeks with less than 1% of the maximum weekly volume), so all series were re-scaled between 0 and 100. These were clustered using hierarchical clustering, computing the pairwise Euclidean distance between words and using Ward’s linkage method (an agglomerative algorithm) to construct the dendrograms shown in 2. clustering was performed in Python, using scipy.cluster.hierarchy.dendrogram^66^. The number of clusters was determined through visual inspection of the dendrogram. This task was performed using data from the pandemic period, which for H1N1 pandemic was between March 2009 and August 2010, and for Covid-19 from December 2019 to September 2020.

#### Modeling and Evaluation

The datasets for seasonal flu were collected similarly to those of the pandemic. They are aggregated by week and seasons were defined by visual inspection, varying from season to season, over the 9 years of data. Each dataset (cases and search time series) in each season was standardized so its mean value was 0 and its standard deviation was 1. The model was trained with 3 seasons and tested with the 4^th^. In the case of Covid-19 in Spain, the data was split around the week with the fewest number of cases (June). The first wave was then used to train and the second to test.

#### Linear Regression

In each case, a model of the form

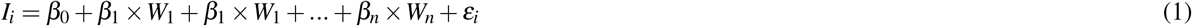

was trained, where *I*_*i*_ is the number of infections in week *i, β*_0_ is the intercept, *β*_1_ to *β*_*n*_ are the coefficients of each search term and *ε*_*i*_ is the error. The coefficients were estimated as to minimize the sum of the square of the errors across all weeks. the regression was implemented in Python using sklearn.linear_model.LinearRegression^67^ with default parameters.

#### Random Forest

For each dataset, a random forest model was trained using sklearn.ensemble.RandomForestRegressor^68^ implemented in Python. The hyperparameters - number of estimators, max features and max depth - were selected through cross validation using *GridSearchCV* from [10,20,50,100,200,500,1000], [0.6,0.8,“auto”,“;sqrt”] and [2,4,5,6] respectively.

## Data Availability

All data used in this manuscript is publicly available.

## Acknowledgments

The authors would like to thank members of the SPAC lab for comments and critical reading of the manuscript. This work was partially funded by FCT grant DSAIPA/AI/0087/2018 to JGS and by PhD fellowships SFRH/BD/139322/2018 and 2020.10157.BD to CHV and SM, respectively.

## Author contributions statement

All authors participated in project conception, data analysis, and paper writing.

## Additional information

