## Supplemental document for "Learning from pandemics: using extraordinary events can improve disease now-casting models"

January 18, 2021

**4 Granger causality** **6**

### 1. FULL LIST OF SEARCH TERMS FOR FLU US AND COVID-19 SPAIN

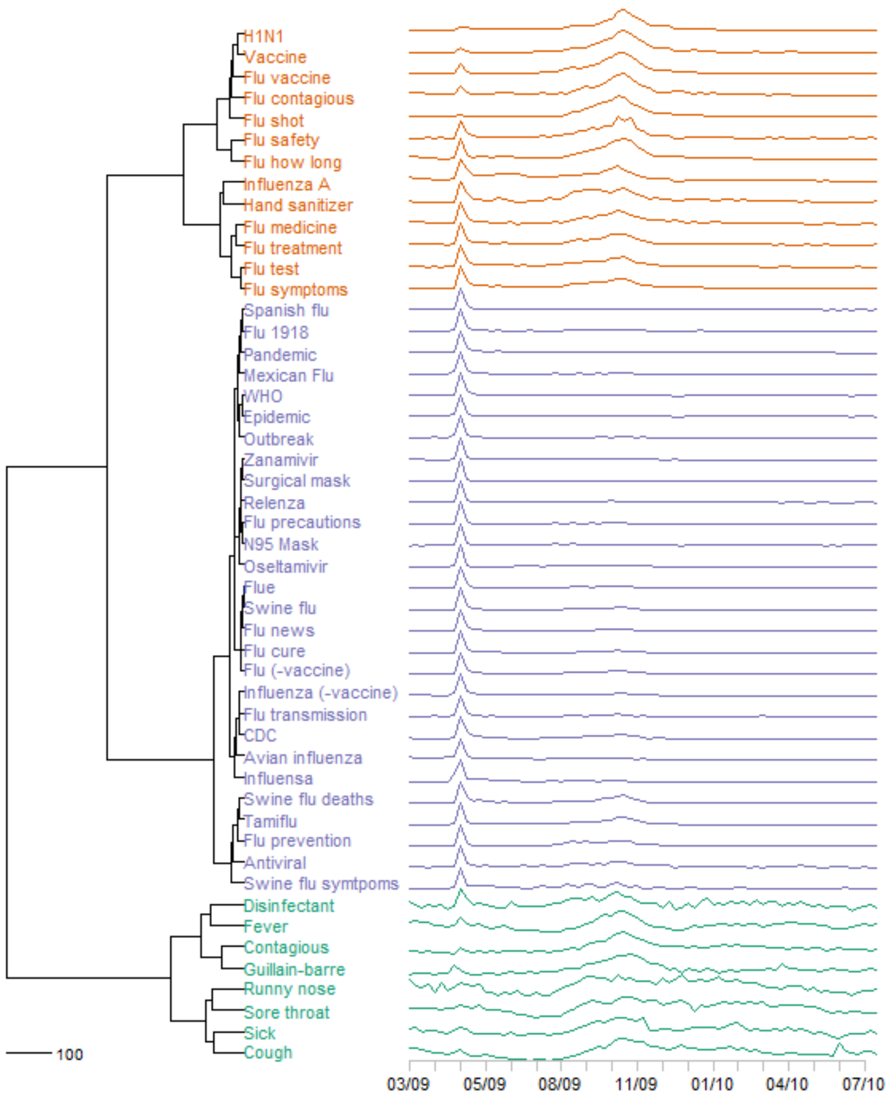

**Fig. S1. Google trends search terms dendogram for US.** The dendrogram was obtained through agglomerative hierarchical clustering using Euclidean distance and Ward's linkage criterion. The pandemic period (March 2009-July 2010) was used and three distinct clusters were identified: Cluster 1 (orange), Cluster 2 (purple), Cluster 3 (green).

**Collected terms US - Flu pandemic:**
H1N1, Vaccine, Flu vaccine, Flu contagious, Flu shot, Flu safety, Flu how long, Influenza A, Hand sanitizer, Flu medicine, Flu treatment, Flu test, Flu symptoms, Spanish Flu, Flu 1918, Pandemic, Mexican Flu, WHO, Epidemic, Outbreak, Zanamivir, Surgical mask, Relenza, Flu precautions, N95 mask, Oseltamivir, Flue, Swine flu, Flu news, Flu cure, Flu (-vaccine), Influenza (-vaccine), Flu transmission, CDC, Avian influenza, Influensa, Swine flu deaths, Tamiflu, Flu Prevention, Antiviral, Swine flu symptoms, Disinfectant, Fever, Contagious, Guillain-barre, Runny nose, Sore throat, Sick, Cough

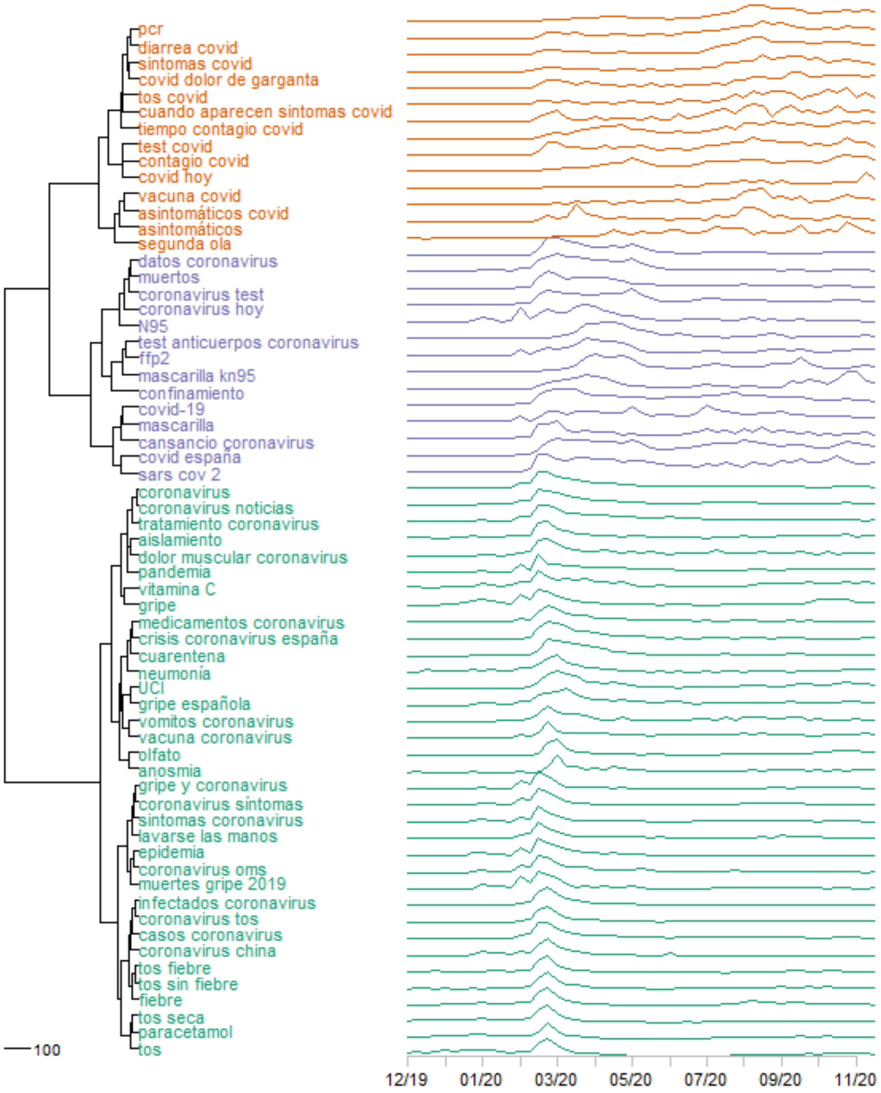

**Fig. S2. Collected terms Spain(ES) dendrogram.** The dendrogram was obtained through agglomerative hierarchical clustering using Euclidean distance and Ward's linkage criterion. The selected period to build the clusters was between December 2019 to September 2020. Three clusters were identified and search terms membership can be observed: Cluster 1 (orange), Cluster 2 (purple) and Cluster 3 (green).

**Collected terms ES - Covid-19 pandemic:**
pcr, diarrea covid, sintomas covid, covid dolor de garganta, tos covid, cuando aparecen sintomas covid, tiempo contagio covid, test covid, contagio covid, covid hoy, vacuna covid, asintomáticos covid, asintomáticos, segunda ola, datos coronavirus, muertos, coronavirus test, coronavirus hoy, N95, test anticuerpos coronavirus, ffp2, mascarilla kn95, confinamiento, covid-19, mascarilla, cansancio coronavirus, covid españa, sars cov 2, coronavirus, coronavirus noticias, tratamiento coronavirus, aislamiento, dolor muscular coronavirus, pandemia, vitamina C, gripe, medicamentos coronavirus, crisis coronavirus españa, cuarentena, neumonía, UCI, gripe española, vomitos coronavirus, vacuna coronavirus, olfato, anosmia, gripe y coronavirus, coronavirus síntomas, sintomas coronavirus, lavarse las manos, epidemia, coronavirus oms, muertes gripe 2019, infectados coronavirus, coronavirus tos, casos coronavirus, coronavirus china, tos fiebre, tos sin fiebre, fiebre, tos seca, paracetamol, tos

### 2. MODEL RESULTS FOR FLU US AND COVID-19 SPAIN

| | Flu | | | | Covid-19 | | | |
|---|---|---|---|---|---|---|---|---|
| | L. Regression | | Random Forest | | L. Regression | | Random Forest | |
| | $R^2$ | RMSE | $R^2$ | RMSE | $R^2$ | RMSE | $R^2$ | RMSE |
| Cluster 1 | 0.83 | 0.17 | 0.86 | 0.14 | 0.96 | 0.04 | 0.84 | 0.16 |
| Cluster 2 | 0.76 | 0.25 | 0.82 | 0.18 | 0.70 | 0.30 | 0.20 | 0.80 |
| Cluster 3 | 0.50 | 0.50 | 0.53 | 0.47 | 0.55 | 0.45 | 0.44 | 0.56 |
| All data | 0.72 | 0.28 | 0.81 | 0.19 | 0.46 | 0.54 | 0.35 | 0.65 |

We collected weekly counts of laboratory-confirmed cases of all strains of flu combined for USA.

### 3. COVID-19 IN THE USA

#### A. Cases and media mention for Covid-19 in the USA

Data from Google search trends (GT), news media and Covid-19 infection cases for the USA was extracted from January 26th to November 15th 2020. We used GT API to obtain the weekly search terms volume, news media was extracted using Media Cloud [1] and the infection cases collected using the Covid-19 tracking API from *The Atlantic*[2].

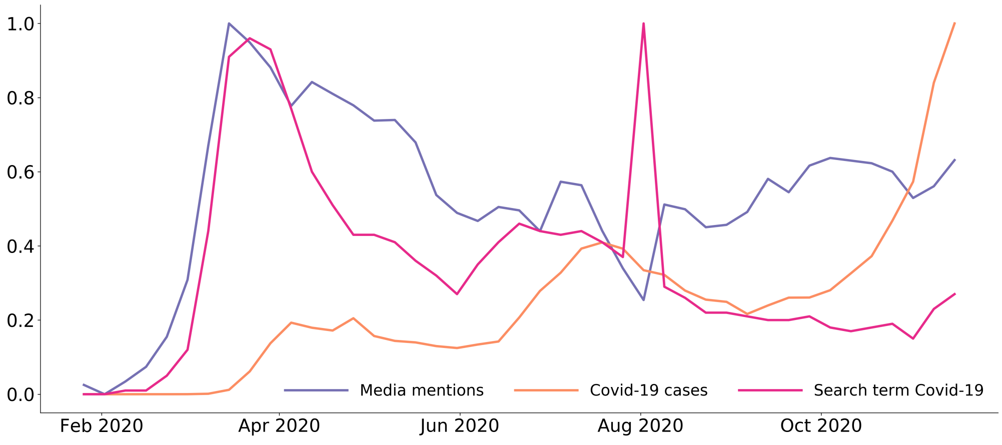

**Fig. S3. Covid-19 cases during the 2020 pandemic in the US.** Weekly cases of Covid-19 (in orage), media mentions (in purple) and the search volume for the search term 'Covid-19' in the United States of America from January 2020 until mid November 2020. We can see a quick increase in media activity that precedes the number of cases of infection. Also, some search terms, like 'Covid-19', have very similar apparent trends to media activity.

#### B. Search Terms

**Collected terms US - Covid-19 pandemic:**

covid-19 testing, covid-19 symptoms, hot spots, covid pneumonia, covid cough, chest pain covid, covid symptoms, covid, loss taste, covid cold symptoms, vaccine, pcr, pcr test, flu shot, stay home, anosmia, when will quarantine end, nk95, contact tracing, covid-19 news, icu beds, icu, loss taste and smell, loss of smell, covid anosmia, quarantine, isolation, coronavirus cases, covid-19 map, coronavirus covid-19, pandemic, coronavirus deaths, spanish flu, flu 2019, flu deaths, deaths, ffp2, covid virus, covid-19 cases, chest pain, nhs, corona, dry cough, coronavirus, cdc, outbreak, epidemic, coronavirus symptoms, sore throat, fever, cold symptoms, coronavirus symptoms vs cold,

cold flu symptoms, flu symptoms, influenza, pneumonia symptoms, cough and fever, respiratory infection, pneumonia, cough, flu, oseltamivir tamiflu, oseltamivir phosphate, rsv, oseltamivir, tamiflu, symptoms influenza, bronchitis contagious, robitussin, bronchitis pneumonia, bronchitis

### C. Clusters

We chose to identify the search terms that best characterize search behavior across the United States. This selection was used to predict new cases across the country. To better understand whether we could also predict new infections locally, we selected three states that had two well-defined waves of infections (train with one wave to test on the second wave). However, the search terms included in each state were not always the same, as terms containing zeros were discarded.

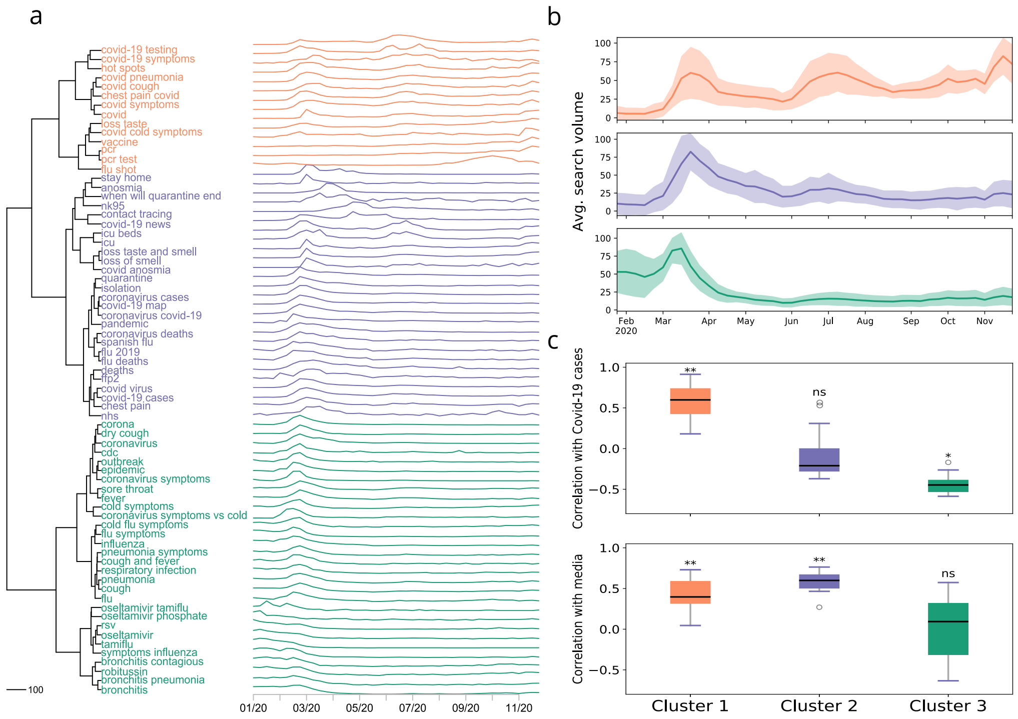

**Fig. S4. Different patterns of searches during the Covid-19 pandemic from January to November 2020 in the USA. a -** Dendrogram summarizing the hierarchical clustering of Google Trends search terms for the Covid-19 pandemic in US. Three clusters are very salient. **b -** Centroid and standard deviation over time for each cluster. The cluster colors correspond to the clusters in a. **c -** Pearson correlation between the cluster centroid and either the Covid-19 cases (top) or the media mentions (bottom).* denotes 0.01 < *p-value* < 0.05, ** denotes *p-value* < 0.001, and *ns* a non significant *p-value*.

### D. Nowcasting

Models were trained with the first wave of Covid-19 cases for all the USA, and the second wave was used to test model predictions. From the results shown in Table S1 we can see that better predictions were obtained with Cluster 1, the one more correlated with cases, in both models.

| Covid-19 USA | | | | |
|---|---|---|---|---|
| | Random Forest | | Linear Regression | |
| | $R^2$ | RMSE | $R^2$ | RMSE |
| All data | 0.63 | 0.37 | 0.35 | 0.65 |
| Cluster 1 | 0.85 | 0.15 | 0.87 | 0.13 |
| Cluster 2 | -0.6 | 1.06 | -25.29 | 26.29 |
| Cluster 3 | -0.46 | 1.46 | -130.99 | 131.99 |

**Table S1.** Predictions results obtained using linear regression and random forest models for the Covid-19 pandemic in the USA.

### 4. GRANGER CAUSALITY

To test whether there was evidence of causality between media, the news and each of the clusters, a lagged regression model was fitted to the data:

$$C(t) = \sum_{\tau=1}^{L} b_\tau \times x_i(t+\tau) + \epsilon_i \tag{S1}$$

where C(t) are the autoregressed values of the cluster centroid at week $t$ (autoregression was performed using the function *diff* in R [3] with order 1), $b_\tau$ are the fitted regression coefficients, $x_i(t)$ are the values of the time series to be tested at week $t$ and $\epsilon_i$ are the residuals. $L$ is the number of lags tested, which was chosen as the one explaining more variance in $C$, using the function *VARselect* [4]. Lags up to four weeks were tried. Granger causality was tested by computing $G = \log(\frac{\sigma_j}{\sigma_i})$, where $\sigma_j$ and $\sigma_i$ are the variance of the residuals in the autoregression and in the lagged regression, respectively. $G$ is expected to follow a $\chi^2$ distribution with $L$ degrees of freedom. The test was implemented using *grangertest* [5].

| Flu | | | | | | |
|---|---|---|---|---|---|---|
| | Cases | | | Media | | |
| | Order | $G$ statistic | $p$ value | Order | $G$ statistic | $p$ value |
| Cluster 1 | 2 | 1.95 | 0.15 | 4 | 1.94 | 0.11 |
| Cluster 2 | 3 | 0.52 | 0.67 | 4 | 11.95 | $\mathbf{3 \times 10^{-7}}$ |
| Cluster 3 | 2 | 1.91 | 0.16 | 45 | 1.21 | 0.31 |
| Covid-19 Spain | | | | | | |
| | Cases | | | Media | | |
| | Order | $G$ statistic | $p$ value | Order | $G$ statistic | $p$ value |
| Cluster 1 | 1 | 0.03 | 0.86 | 1 | 0.15 | 0.70 |
| Cluster 2 | 1 | 0.95 | 0.34 | 1 | 0.03 | 0.86 |
| Cluster 3 | 2 | 0.76 | 0.48 | 1 | 0.79 | 0.38 |

**Table S2.** Results of a Granger-causality test on whether confirmed cases (left-side columns) or media (right side columns) preceded the centroids of each cluster. We only find evidence of Granger-causality in the case of flu media mentions and cluster 2. In the table, "Order" represents the number of lagged weeks.
